# Pharmacokinetic effects of a single-dose nutritional ketone ester supplement on brain glucose and ketone metabolism in alcohol use disorder

**DOI:** 10.1101/2023.09.25.23296090

**Authors:** Xinyi Li, Anthony J. Young, Zhenhao Shi, Juliana Byanyima, Sianneh Vesslee, Rishika Reddy, Timothy Pond, Mark Elliott, Ravinder Reddy, Robert K. Doot, Jan-Willem van der Veen, Henry R. Kranzler, Ravi Prakash Reddy Nanga, Jacob G. Dubroff, Corinde E. Wiers

## Abstract

Acute alcohol intake decreases brain glucose metabolism and increases brain uptake of acetate, a metabolite of alcohol. This shift in energy utilization persists beyond acute intoxication in individuals with alcohol use disorder (AUD), and may contribute to alcohol craving. We recently found that ketone therapies decrease alcohol withdrawal and alcohol craving in AUD. Here, we studied the effects of a single-dose ketone ester (KE) supplement on brain energy metabolism and alcohol craving. Five AUD and five healthy control (HC) participants underwent two ^18^F-fluorodeoxyglucose positron emission tomography (PET) scans, after consumption of 395 mg/kg KE or without (baseline), in randomized order. In the AUD group, KE reduced alcohol craving scores compared to baseline. KE decreased blood glucose levels and elevated blood β-hydroxybutyrate (BHB) levels compared to baseline in both groups. Whole-brain voxel-wise maps of the cerebral metabolic rate of glucose (CMRglc) decreased by 17% in both groups, with the largest KE-induced CMRglc reductions in the frontal, occipital, and cingulate cortices, hippocampus, amygdala, and insula. There were no group differences between AUD and HC in blood or FDG measures, and no correlations between reductions in craving with CMRglc. Cingulate BHB levels, as assessed with ^1^H-magnetic resonance spectroscopy in 5 participant with AUD, increased 3-fold with KE compared to baseleline. In sum, administration of a single dose of KE rapidly shifted brain energetics from glucose to ketone metabolism in HC and AUD. KE also reduced ratings of alcohol craving, demonstrating its potential clinical effectiveness for supporting brain health and alcohol craving in AUD.

## 1 Introduction

Alcohol use disorder (AUD) is a chronic, relapsing condition that is associated with a substantially elevated risk for neurocognitive impairments, co-occurring medical and psychiatric disorders, and mortality [1, 2]. In the United States, approximately 95,000 alcohol-related deaths were reported annually between 2011 and 2015, making alcohol the third-leading cause of preventable death [1]. However, only 1.6 % of patients with AUD receive FDA-approved medications to treat the disorder [3], and treatment efficacy varies among individuals [4].

Improving brain energetics provides a promising novel approach to AUD treatment [5, 6]. Alcohol intake acutely increases brain utilization of acetate, a metabolite of alcohol, at the expense of glucose [7, 8]. In individuals with AUD, low brain glucose and high acetate metabolism persist beyond acute intoxication [8–10], and the lack of acetate as an energy source during abstinence may contribute to alcohol withdrawal signs and symptoms, alcohol craving, and relapse in individuals with AUD [5]. Ketone bodies (β-hydroxybutyrate [BHB], acetoacetate, and acetone) resemble acetate and provide an alternative to glucose as an energy source in the brain [11]. Previous work in our laboratory has demonstrated the efficacy of a high-fat, low-carbohydrate ketogenic diet (KD) as an intervention for reducing alcohol withdrawal severity and craving in AUD patients undergoing alcohol detoxification [6, 12]. These findings are concordant with preclinical models showing that the KD reduces both the signs of alcohol withdrawal [13–15] and alcohol self-administration [6, 16].

Given the restrictive nature of the KD and poor patient adherence to the strict dietary regimen [17], exogenous ketone supplementation provides a more feasible alternative to a KD in elevating plasma ketone bodies [18]. Exogenous ketone ester (KE) supplementation is a well-tolerated intervention that may provide various health benefits. For example, KE improved cognition in preclinical and clinical models of Alzheimer’s Disease [19–21], stabilized brain networks—thereby protecting the brain against hypometabolism and premature aging [22]—and decreased appetite in healthy human volunteers [18]. Preclinical studies of AUD showed that KE decreases alcohol withdrawal signs, similar to KD [13]. In human volunteers, pairing exogenous ketones with a low dose of alcohol decreased blood and breath alcohol levels and alcohol liking and wanting [23]. Ketone bodies, endogenously produced following adherence to KD, enter the brain through monocarboxylate transporters [24]. However, the ability of exogenous KE to cross the blood-brain barrier and shift brain glucose to ketone metabolism has not been studied.

Here, we tested the pharmacokinetic effects of a single dose of KE on brain glucose metabolism, measured using positron emission tomography (PET) with ^18^F-fluoro-deoxyglucose (^18^F-FDG), in participants with AUD and in non-dependent healthy volunteers. All participants underwent two PET imaging sessions, one at baseline (without KE) and one following the KE intervention. We hypothesized that a single dose of KE would decrease the cerebral metabolic rate of glucose (CMRglc) more in AUD than non-dependent controls, and that KE would reduce ratings of alcohol craving in AUD participants. We additionally investigated KE’s effects on brain BHB levels using ^1^H-MRS in the dorsal anterior cingulate cortex (dACC) in 5 subjects with AUD to examine the degree to which KE crosses the blood-brain barrier. We chose the dACC as the main region of interest for ^1^H-MRS because our previous research showed significant effects of KD on brain ketone and glutamate metabolism and functional reactivity to alcohol cues in that brain region [6, 25].

## 2 Methods

### 2.1 Participants

We recruited participants between the ages of 21 and 65 years of age. Five individuals with AUD and 5 healthy controls (HC) completed the ^18^F-FDG procedures under study protocol NCT05015881. **Table 1** lists the demographics and clinical characteristics of the 10 participants. Individuals were included in the AUD group if they met the DSM-5 criteria for current AUD, and reported having consumed an average of 15 or more standard drinks per week in the last month on the Timeline Follow-back interview [26], greater than a 1-year history of heavy drinking, and at least one drink during the week prior to the study visit. Individuals in the HC group consumed less than 15 total standard drinks and 3 standard drinks per occasion in the last month and scored less than 6 on the Alcohol Use Disorders Identification Test (AUDIT), indicative of non-hazardous drinking behavior [27]. Exclusion criteria for all participants included: 1) a current DSM-5 diagnosis of a major psychiatric disorder (other than AUD in the AUD group) based on medical history and the structured Mini International Neuropsychiatric Interview (MINI), 2) unwilling or unable to refrain from taking psychoactive medication within 24 hours of study participation, 3) history of seizures, 4) HIV seropositivity, 5) a history of head trauma with loss of consciousness for more than 30 min or associated skull fracture/abnormal MRI, 6) any contraindication for MRI (e.g., presence of ferromagnetic objects or claustrophobia), 7) a current, clinically significant physical disease or abnormality detected by medical history, physical examination, or routine laboratory evaluation that could impact brain function, 8) pregnancy or lactation, and 9) a body mass index >35 kg/ m^2^. We also excluded individuals who tested positive for any substance other than cannabis on a urine drug test or had a breath alcohol concentration above 0.00% upon arrival to the study visits. Five individuals with AUD completed the ^1^H-MRS study under protocol NCT0461678 with similar inclusion and exclusion criteria for AUD (see supplementary Table 1 for demographics of the ^1^H-MRS cohort; n=3 overlapped with the PET ^18^F-FDG study). The study protocols were approved by the University of Pennsylvania Institutional Review Board and all participants provided written informed consent before study participation.

**Table 1.** Demographics and clinical characteristics of the participants in the^18^F-FDG PET study. Data are presented as mean ± SD.

| Characteristic | HC (n=5) | AUD (n=5) |
| --- | --- | --- |
| Sex at birth | 2 females/ 3 males | 1 female/ 4 males |
| Age (years) | 36.2 ± 5.3 | 31.0 ± 8.7 |
| Ethnicity | 1 African American/ 3 Caucasian/ 1 Asian | 2 African American/ 3 Caucasian |
| BMI (kg/m<sup>2</sup>) | 23.7 ± 2.6 | 30.5 ± 2.4* |
| Shipley IQ score | 121.8 ± 7.2 | 108.4 ± 21.3 |
| TLFB drinks/week |  |  |
| Screening | 0.7 ± 0.7 | 26.8 ± 9.2* |
| Baseline | 1.1 ± 1.2 | 26.1 ± 11.0* |
| KE | 1.1 ± 1.3 | 24.2 ± 12.7* |
| Days since last drink (30 days) |  |  |
| Baseline | 1.6 ± 0.9 | 9.0 ± 12.1 |
| KE | 2.2 ± 1.5 | 8.0 ± 12.4 |
| AUDIT | 1.8 ± 1.3 | 14.8 ± 4.1* |

\*Indicates significant differences ( $p < 0.05$ ) between HC and AUD. Abbreviations: AUD, alcohol use disorder; AUDIT, Alcohol Use Disorders Identification Test; BMI, body mass index; HC, healthy control; KE, ketone ester; TLFB, Timeline Follow-back interview.

### 2.2 Study design and intervention

Both study protocols used a cross-over design, in which a computer-generated randomization scheme assigned 50% of subjects to receive no KE (baseline) or 395 mg/kg (2.2 mmol/kg) body weight ketone ester (KE, (R)-3-hydroxybutyl (R)-3-hydroxybutyrate, TdeltaS Global Inc.) on study visit 1, and the reverse condition on visit 2. There were no differences in drinking behavior between KE and baseline study days (**Table 1**).

The PET ^18^F-FDG study was open-label, and no intervention was provided at the baseline visit. Participants fasted for at least 6 hours before the study, as is standard for ^18^F-FDG study protocols. During the intervention visit, KE was diluted with diet soda or sparkling water by a trained staff member and administered to participants 1 hr prior to the PET scan. Blood levels of glucose and BHB were assessed at arrival, immediately prior to ^18^F-FDG injection, and at 30 and 60 min post-injection (**Figure 1A**). Measurements were made following finger sticks using commercially available meters (Precision Xtra, Abbott Laboratories, Chicago, IL, USA). Following the PET scan on each of the lab days, AUD patients completed the Alcohol Urge Questionnaire (AUQ), which consists of eight statements about feelings and thoughts about alcohol drinking as when completing the questionnaire (i.e., right now) on a 7-point Likert scale (strongly disagree – strongly agree). Thus, the total score ranges from 7-56 [28].

**Figure 1.**
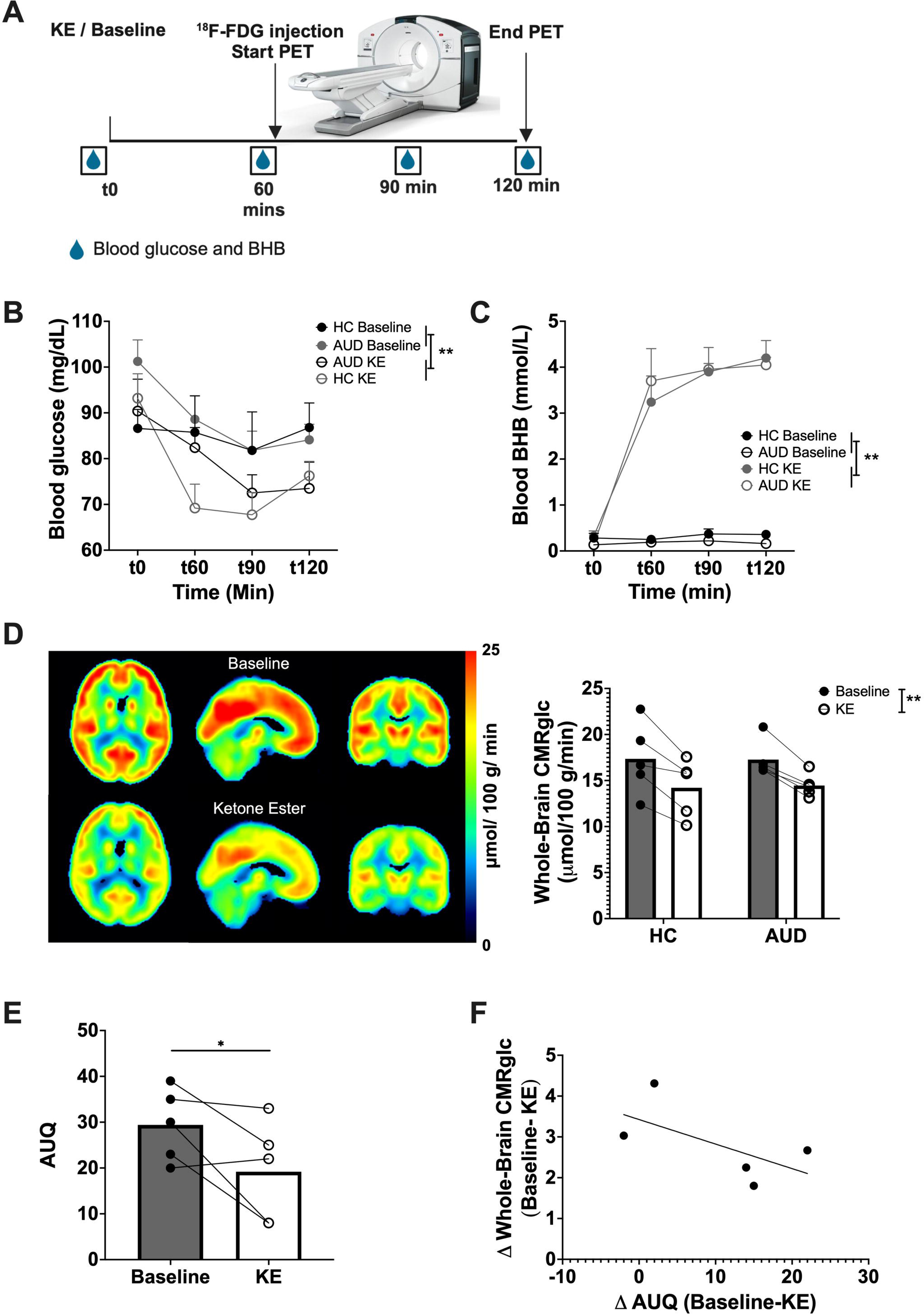
Blood ketone and glucose levels and brain glucose metabolism at baseline and following ketone ester (KE) in healthy individuals and individuals with AUD. **(A)** Timeline of study events on the days of the positron emission tomography scans. Participants arrived at the study center after fasting for at least 6 hours and were administered no intervention (baseline) or the KE ∼1 hr before the scan. **(B)** Blood glucose levels were significantly lower and **(C)** BHB levels were significantly higher following KE than at baseline. **(D)** Average CMRglc maps of healthy individuals and individuals with AUD combined following KE and at baseline. Whole-brain CMRglc decreased with KE *vs.* baseline. **(E)** In individuals with AUD, AUQ scores were lower following KE than at baseline. **(F)** Reductions in AUQ scores (baseline-KE) did not correlate with reductions in whole-brain CMRglc. Mean ±SEM. **Indicates significant differences between baseline and KE, p<0.05. *Indicates significant differences between baseline and KE, p<0.05. Abbreviations: AUD, alcohol use disorder; AUQ, Alcohol Urge Questionnaire; BHB, β-hydroxybutyrate; CMRglc, cerebral metabolic rate for glucose consumption; FDG, fluorodeoxyglucose; KE, ketone ester; PET, positron emission tomography.

The MRI study was double-blinded and placebo-controlled, and a timeline of events is provided in **Figure 2A**. Participants (n=5 individuals with AUD) were instructed to fast overnight, and on the morning of the study visit were provided a low-calorie breakfast ∼2 hours before the scan. They then received KE or placebo 15 min prior to the start of the scan or 45 min prior to the ^1^H-MRS sequence. The study drinks were prepared by the Penn Investigational Drug Service, and an isocaloric dextrose drink that was taste-matched using a bitter additive (Symrise, Inc., Teterboro, NJ, USA) served as the placebo.

**Figure 2:**
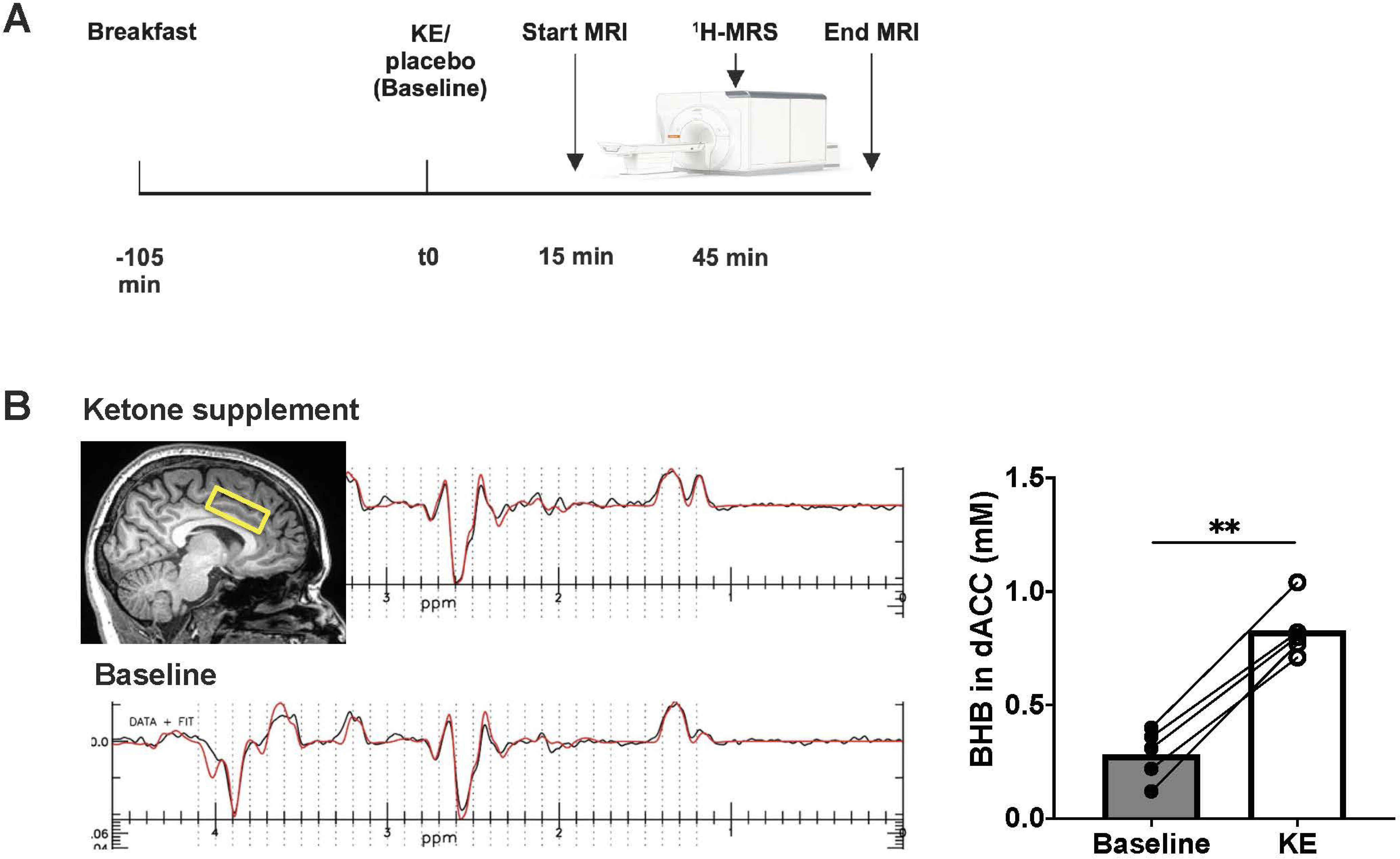
Brain ketone levels at baseline and following ketone ester in individuals with AUD. **(A)** Schematic of the experimental design. Participants arrived at the study center following an overnight fast, were provided a standardized breakfast, and received KE or placebo (baseline). MRI acquisition started 15 min following the administration of the study drinks and included a ^1^H-MRS sequence to measure BHB. **(B)** Voxel for ^1^H-MRS was placed with dACC (40×30×15mm) as the region of interest and BHB was detected in the ^1^H-MRS spectra at 1.19 ppm. Brain BHB levels were significantly higher 45 min after KE intake (mean= 0.83± 0.13 mM) than at the baseline visit (mean= 0.28± 0.11). Mean ±SEM. **Indicates significant differences between baseline and KE, p<0.05. Abbreviations: BHB, β-hydroxybutyrate; dACC, dorsal anterior cingulate cortex, KE, ketone ester.

### 2.3 ^18^F-FDG PET scanning and processing

On two separate study days participants underwent a 60-min ^18^F-FDG PET/CT scan using the PennPET Explorer [29], a 140-cm axial field of view whole-body PET scanner. Venous catheters were placed in the antecubital vein bilaterally for radiotracer injection and blood sampling. Commercially manufactured *^18^F*-FDG (∼10 mCi) was injected intravenously over a period of approximately 1 min. During the PET imaging procedures, the subjects rested quietly under dim illumination and minimal acoustic noise. To ensure that subjects did not fall asleep, they were monitored throughout the procedure and were asked to keep their eyes open.

Dynamic PET images were reconstructed into 4-mm voxels using time-of-flight list-mode ordered-subsets expectation maximization into 39 frames: 12 x 5 s, 6 x 10 s, 3 x 20 s, 2 x 30 s, 6 x 60 s, and 10 x 300 s. Images were corrected for interframe motion and analyzed in PMOD v3.7 (PMOD Technologies, Zurich, Switzerland). Whole brain and voxel-wise flow (K) were computed using Gjedde-Patlak graphical modeling [30], which models tissue uptake of ^18^F-FDG and its subsequent phosphorylation that traps it within the cell. An equilibration time t* of 20 minutes was used for ^18^F-FDG transport to stabilize for fitting [31]. Individual’s blood functions used an image-derived aortic arch blood pool SUV_peak_ input function with a plasma:whole blood ratio of 1:1. Measured ^18^F-FDG flow was converted to glucose metabolism using the formula MR_Glc_ = K*PG/LC, where PG is the averaged plasma glucose levels from pre-injection, 30-min, and 60-min post-injection samples, and LC is a lumped constant of 0.8 [32]. The cerebral metabolic rate for glucose consumption (CMRglc) maps in μmol/100 mL/min were aligned to the subject’s structural MRI and then normalized to the MNI template with a 2-mm isotropic resolution using the FSL Software Library (version 5.0; http://www.fmrib.ox.ac.uk/fsl)[33].

### 2.4 Brain MRI imaging and processing

Participants underwent an MRI on a 3.0T Magnetom Prisma scanner (Siemens Medical Solutions USA Inc., Malvern, PA) equipped with a 64-channel head coil. Structural images were acquired using T_1_-weighted magnetization-prepared rapid-acquisition gradient-echo (MPRAGE) sequences with the following parameters: TR 2400 ms; TE 2.24 ms; TI 1060 ms; FOV 256 mm; 208 slices; slice thickness 0.8 mm; flip angle 9°; effective voxel resolution, 0.8 x 0.8 x 0.8 mm.

Localized proton magnetic resonance spectroscopy (^1^H-MRS) was performed in a region of interest (ROI) located in the dACC (40×30×15mm), for which we previously demonstrated a sensitivity to ketones [34, 35]. A semi-Laser (sLaser) spectral editing sequence for BHB was acquired, using an adapted sequence previously described [36]. The parameters were as follows: TR 2000 ms; TE 72 ms; 256 averages. Editing was performed with a Gaussian pulse of 18.5 ms, amplitude of 47 Hz, and a bandwidth of 80 Hz at FWHM. For edit-on, the pulse was centered at 4.14 ppm while for edit-off scans, it was centered at 5.26 ppm relative to a water position of 4.70 ppm. First- and second-order shimming was used to maximize the magnetic field B_0_ homogeneity in the voxel. Spectral fitting of the ^1^H-MRS datasets was carried out with in-house written fitting software in IDL (NV5Geospatial, Broomfield, CO, USA) [37] with a modified basis set for the specific editing sequence and for ketone bodies [38]. The basis set was simulated using GAMMA, part of the VESPA package [39]. The results from the analysis were inspected for nonrandom residuals and baseline fitting. Spectra with signal-to-noise ratio (SNR) < 15 and linewidth > 0.1 ppm were excluded from further analyses. A Cramér–Rao lower bound (CRLBs) of 20% for each individual peak was used as a quality criterion [37]. To place the voxel in the same dACC ROI during the second scan, we used the program ImScribe (https://www.med.upenn.edu/CAMIPM/imscribe.html), which is designed to allow reproducible selection of the same anatomical ROI in Siemens MRI studies within the same subject over multiple MRI scans. ImScribe uses high-resolution T_1_-weighted images and the spectroscopy voxel information from the first scan as target template and T_1_-weighted images of the subsequent scan and performs affine coregistration, which provides information on the new spectroscopic voxel placement for the later scans [40, 41]. This was completed (∼2 min) and the ROI was transferred to the Siemens console as a DICOM image and used to prescribe the ^1^H-MRS acquisition.

### 2.5 Statistical analyses

Statistical analyses on blood glucose and BHB concentrations were conducted using SPSS (IBM, Armonk, NY). We performed linear mixed-effects analysis to examine the effects of the intervention (KE vs. baseline) x group (AUD vs. HC) x time (4 time points per session) on blood glucose and BHB concentrations. The models also included subject-specific random intercepts. *Post hoc* pairwise comparisons were performed with Bonferroni correction. A similar linear mixed-effects model was used to examine the effects of the intervention x group on whole-brain CMRgluc. Within the AUD participants, the effect of the intervention on AUQ sum scores and cingulate BHB levels was analyzed with paired t-tests (one-tailed p<0.05).

Voxel-wise comparisons were performed in SPM12 (http://www.fil.ion.ucl.ac.uk/spm/) using a mixed design ANOVA with Intervention (KE vs baseline) as within-subject and Group (AUD vs HC) as between-subject factors at a cluster-defining threshold of uncorrected p<0.0001 and cluster size k>100, and cluster-level threshold of familywise error (FWE)-corrected p<0.05. Exploratory Pearson’s correlations were performed on the associations between AUQ craving and CMRglc at baseline and KE, and on the delta baseline-KE AUQ and CMRglc measures.

## 3 Results

### 3.1 Blood glucose and BHB levels

Blood glucose levels were lower during the KE intervention than at baseline (main effect of Intervention: F_1,50.9_=18.6, p<0.001) and decreased with Time (main effect of time: F_3,51.0_=12.7, p<0.001), but there were no significant effects of Group, or interaction effects (p’s> 0.05) (**Figure 1B**). BHB levels were significantly higher following KE than at baseline (Intervention: F_1,49.5_=465.8, p<0.001; Intervention x Time: F_3,49.0_=46.8, p<0.001) and increased with Time (F_3,49.5_=51.6, p<0.001), but there were no effects of Group or any of its interactions (p’s> 0.05). *Post hoc* analysis showed significant differences in BHB levels between baseline and KE at 60, 90, and 120 min post-KE administration. KE increased blood BHB levels approximately 20-fold from a pre-KE level of 0.2± 0.2 mM to 4.1± 1.3 mM 120 min following administration. (**Figure 1C**).

### 3.2 Brain CMRglc (PET FDG)

There was a main effect of Intervention on whole-brain CMRglc, which decreased by 17% with KE (mean CMRglc = 14.3± 2.3 µmol/100g/min ) *vs.* baseline (mean = 17.3± 2.9 µmol/100g/min) (F_1,8_=49.1, p< 0.001, Cohen’s d = 2.3). There was no significant main effect of Group (F_1,8_=0.002, p=0.97), and no significant Intervention x Group interaction (F_1,16_=0.2, p=0.70) (**Figure 1D**). Model fits for a representative patient at baseline and after KE are provided in **Supplemental Figure 1**. Voxel-wise whole-brain analysis revealed widespread CMRglc reductions by KE *vs.* Baseline, with the largest KE-induced CMRglc reductions in the frontal cortex, occipital cortex, cingulate cortex, insula, hippocampus, and amygdala (all p_FWE_<0.05) (**Figure 1D**, **Table 2**).

**Table 2.** Brain regions demonstrating a significant main effect of Intervention on CMRglc (PET with ^18^F-FDG) using Statistical Parametric Mapping with a cluster-defining threshold of uncorrected p<0.0001, cluster size k>100, and cluster-level threshold of familywise error (FWE)-corrected p<0.05.

| Brain Region | Cluster size (# of voxels) | Z | MNI Coordinates |
| --- | --- | --- | --- |
| **Baseline > KE contrasts** |  |  |  |
| L Middle frontal gyrus | 390 | 5.57 | -32, 24, 34 |
| L Lingual gyrus | 5066 | 5.26 | -20, -68, -6 |
| L Cuneus | 313 | 5.01 | -20, -84, 24 |
| R Middle occipital gyrus | 809 | 4.99 | 26, -96, 6 |
| L Medial frontal gyrus, cingulate gyrus | 1387 | 4.97 | -8, -8, 52 |
| R Pons | 730 | 4.92 | 8, -28, -38 |
| R Superior temporal gyrus | 585 | 4.84 | 70, -20, 6 |
| R Insula | 180 | 4.81 | 40, 0, 18 |
| L Middle and inferior temporal gyrus | 135 | 4.80 | -58, 0, -32 |
| R Middle temporal gyrus | 130 | 4.76 | 52, -32, -10 |
| R Superior and Inferior parietal lobule | 303 | 4.70 | 28, -56, 48 |
| R Inferior frontal gyrus | 149 | 4.70 | 32, 28, 28 |
| R Precuneus | 353 | 4.64 | 8, -60, 60 |
| R Hippocampus, amygdala | 526 | 4.57 | 34, -4, -24 |
| R Cingulate gyrus | 416 | 4.56 | 10, 10, 34 |
| R Fusiform gyrus | 440 | 4.38 | 32, -56, -10 |
| R Middle occipital gyrus | 115 | 4.27 | 48, -70, 0 |
| **Baseline < KE contrasts** |  |  |  |
| No significant clusters |  |  |  |
| Abbreviations: KE, ketone ester; L, left; MNI, Montreal Neurological Institute; R, right |  |  |  |

### 3.3 Alcohol craving

In individuals with AUD, KE decreased AUQ craving levels by an average of 34.0% ± 36.3 (t_4_=2.3, p= 0.04, Cohens’ d=1.1) (**Figure 1E**). Supplemental Table 2 lists the means of the AUQ sum score and individual questions for KE and baseline. KE-induced reductions in CMRglc did not covary with reductions in AUQ craving (r= -0.62, p= 0.26) (**Figure 1F**). Nevertheless, whole-brain CMRglc correlated with AUQ when both interventions were pooled together (r=0.68, p=0.03), such that lower AUQ scores were associated with lower whole-brain CMRglc (**Supplemental Figure 1S**).

### 3.4 Brain BHB (^1^H-MRS)

BHB levels in the dACC were significantly higher 45 min after KE intake (mean= 0.83± 0.13 mM) than at baseline (mean= 0.28± 0.11 mM) (t_4_=13.4, p<0.001, Cohen’s d=5.9), as measured with ^1^H-MRS in 5 AUD participants (**Figure 2**).

## 4 Discussion

We found that a single dose of KE rapidly shifted brain glucose to ketone metabolism in human volunteers either with or without AUD. The^18^F-FDG PET findings revealed a 17% reduction in whole brain CMRglc within 1-2 hr after oral intake of KE, and voxel-wise analyses demonstrated significant decreases in the frontal, occipital, and cingulate cortices, hippocampus, amygdala, and insula. KE also increased cingulate BHB levels 3-fold, and reduced alcohol craving ratings in individuals with AUD.

The brain energetic switch from glucose to ketones following a 0.395 g/kg oral KE dose appeared in brain regions similar to those reported by Volkow et al. [7] for a brain energetic switch from glucose to acetate following oral administration of alcohol at a dosage of 0.75 g/kg. Volkow et al. found the greatest reductions in glucose metabolism in the occipital cortex and cerebellum (both 16% reductions), while brain acetate levels showed the greatest increases in the same brain areas. Despite our hypothesis that reductions in CMRglc would be greater in AUD than controls, as was found for brain acetate levels in response to alcohol among occasional social drinkers and heavy drinkers [7], we did not detect group differences in CMRglc following KE. The KE-induced reductions in alcohol craving support our hypothesis that ketones, resembling acetate metabolism, reduce alcohol craving and consumption by supporting brain energy. The reduction in alcohol craving with KE is also in line with reductions in alcohol wanting and liking after KE and alcohol consumption in healthy volunteers [23], reductions in brain craving responses to alcohol cues in AUD patients on a KD [6, 12] and reductions in alcohol consumption in alcohol-dependent rodents on a KD [6, 42].

Glucose is the primary fuel for the brain and is necessary for normal neural functioning, including the maintenance of the resting potential, generation of an action potential, and synthesis of neurotransmitters and neuromodulators [43, 44]. When glucose is not readily available, such as during fasting or the adherence to a low-carbohydrate, high-fat KD, ketone bodies are produced endogenously in the liver, carried through the bloodstream, and transported into neurons by monocarboxylate transporters, where they are utilized for fuel [5]. Several exogenous KE supplements are commercially available and provide a faster, less dietary restrictive alternative to a KD in achieving nutritional ketosis [18, 22, 45]. As demonstrated here, KE elevated blood BHB levels within 1-2 hr of administration to ∼4.1 mM, a level corresponding to 1-2 wk of adherence to a KD [34]. The blood BHB levels in our study are congruent with those previously reported with the same dose of KE [45], which also reported reductions in appetite.

Localized ^1^H-MRS provides a non-invasive method to measure ketone levels in the brains and has been used in individuals adhering to a KD [38, 46]. Using ^1^H-MRS, we demonstrated a rapid and efficient brain uptake of BHB following KE administration and significantly higher BHB levels in the dACC during the KE session compared to the baseline session. Additionally, our findings indicated that KE reduced blood glucose levels, consistent with findings from other studies [47], and may be attributable to a KE-induced increase in insulin levels [45, 47, 48].

Using dynamic PET, we found an average 17% reduction in CMRglc from baseline to KE, comparable to that previously observed following 4 days of a KD intervention [11]. It is possible that the lower glucose and increased insulin in blood for our cohort during the KE session also increased the lumped constant used in Gjedde-Patlak modeling of ^18^F-FDG PET, leading to a potential overestimation of CMRglc [49, 50]. However, it is important to note that reductions in CMRglc have been found to be proportionate to plasma BHB levels [51], and not to hypoglycemia *per se* [11]. In a previous study, plasma glucose levels correlated inversely with brain glucose metabolism and hyperglycemia was associated with significantly lower brain glucose metabolism [52]. However, a previous PET study showed that a decrease in CMRglc in individuals following adherence to a KD correlated with an increased cerebral metabolic rate of acetoacetate [11]. Therefore, the reduction in CMRglc we observed following KE administration likely reflects a shift in energy utilization from glucose to ketones. However, dual tracer PET scanning with FDG and ketone tracers such as [^11^C]acetaoacetate are required to assess dynamic scanning of both brain ketone and glucose metabolism in the same scanning session.

Emerging evidence attributes heavy alcohol consumption and AUD to deficiencies in brain energy metabolism [5, 53]. Acetate is a byproduct of alcohol metabolism and produces energy through conversion to acetyl-coenzyme A and the metabolism in the tricarboxylic acid (TCA) cycle [54]. Alcohol intoxication acutely increases plasma levels of acetate and brain uptake of acetate, and decreases brain glucose utilization [53, 55]. Chronic alcohol consumption results in metabolic adaptations that exacerbate the shift in brain energetics seen with acute intoxication. Thus, heavy drinkers demonstrate higher levels of ^13^C-labeled brain acetate [9] and lower glucose metabolism [53, 56, 57] than light drinkers. Similarly, proteomic analysis in post-mortem human brain tissues and neurometabolic analysis in rodents shows a reduction in glucose metabolic pathways (i.e., glycolysis and gluconeogenesis) and increased expression of monocarboxylate transporters after chronic alcohol use [58, 59]. It has been hypothesized that the deprivation of acetate as an energy source and the inability to abruptly transition to utilizing glucose during abstinence contributes to alcohol withdrawal signs and symptoms and alcohol craving in individuals with AUD undergoing detoxification [5, 53]. Ketone bodies are similar to acetate in that they are converted to acetyl-coenzyme A and produce energy in the TCA cycle. Targeting brain energetics with a KD intervention was previously shown to increase blood and brain ketone levels and attenuate withdrawal signs and symptoms and alcohol craving in individuals with AUD undergoing alcohol detoxification treatment [34]. Here, we demonstrated that KE rapidly increases blood and brain BHB levels and reduces the need for brain glucose utilization in individuals with AUD, and may thus serve as a more palatable alternative to a KD intervention in treating individuals with AUD.

Our study has several limitations and identifies potential areas for future study. First, although we employed a cross-over design to mitigate the potentially confounding effects of inter-individual differences, the study sample is small. Second, the non-significant difference between healthy controls and individuals with AUD on brain glucose metabolism that we observed, contrary to that previously reported, may be attributable to a lower level of alcohol consumption in our AUD subjects (3.8 standard drinks/day) compared to previous studies (8.6-10.6 standard drinks/day) [53, 57, 60, 61]. Furthermore, brain levels of BHB were only assessed in a separate cohort of individuals with AUD and not in healthy controls. Thus, future studies are needed to compare KE-induced increases in BHB levels between individuals with AUD and non-dependent volunteers and to ascertain whether KE-induced elevations in ketones and reductions in CMRglc are greater among individuals who are consume high levels of alcohol. Third, because the measures were assessed in a separate cohort of participants, we were unable to correlate brain BHB levels with CMRglc. However, in a previous study in healthy participants maintained on a KD, cerebral metabolic rates of acetoacetate and glucose were highly correlated (r=-0.91, p< 0.001) [11]. It is likely that, similarly to KD, exogenous KE elevates blood BHB, which subsequently enters the brain and induces a shift in brain energetics. Also, small differences in the experimental design of the PET and MRI studies may have confounded the comparison of the effects of KE on brain glucose and BHB. Participants were provided a standardized meal before the MRI and remained fasting as per the PET protocol. At the baseline visits, a placebo was administered in the MRI study, whereas no intervention was provided in the PET study. Fourth, ^1^H-MRS cannot differentiate between intracellular and extracellular levels of BHB and is thus limited in assessing BHB utilization in the brain. Fifth, *post hoc* pairwise analysis showed lower blood glucose in the KE treatment arm than the baseline visit prior to KE administration, which may have been confounding. However, all CMRglc analyses were corrected for blood glucose levels and likely did not affect the strong KE-induced reductions in CMRglc. Indeed, previous studies reported an inverse relationship between blood and brain glucose, such that higher brain glucose uptake was seen in individuals with lower blood glucose levels [52, 62, 63]. Therefore, our findings of decreased brain glucose uptake with KE *despite* lower blood glucose levels appear to reflect the robust effect of KE in altering brain energetics. Last, the implications of KE-induced changes in brain energetics on clinical outcomes in AUD are unknown. To address this gap in the literature, we plan to conduct studies to examine the effects of KE on alcohol withdrawal and alcohol craving in individuals with AUD.

In sum, our findings provide the first evidence that KE administration rapidly shifts brain energetics from glucose to ketone metabolism, in both healthy volunteers and participants with AUD, and reduces alcohol craving in those with AUD. Understanding the neurometabolic effects of KE may provide a scientific basis for studies examining the feasibility of using KE to treat AUD, or other neuropsychiatric and neurological disorders. Such findings may also help to identify potential neural mechanisms for these clinical effects.

## Data Availability

All data produced in the present study are available upon reasonable request to the authors

## Acknowledgments

We thank Laurie Downing, Kim Olson, Klaudia Glogowska, Rachel Weyl, Erin Schubert, Matt Furey, Regan Scheffer, Kayla Spooner, Reagan Wetherill, Nora Volkow, Paco Bravo, Aiden Adams, David Mankoff, Robert Mach, and the Penn PET Center for their contributions.

